## Supplemental Data 1 for "Data-driven methodology for discovery and response to pulmonary symptomology in hypertension through AI and machine learning: Application to COVID-19 related pharmacovigilance" (from another version of this preprint: version 1, 2021-06-12)

**Running title:** AI and machine learning data-driven methods: COVID related pharmacovigilance

Table S1. Contribution of Pulmonary ADEs in 2D and 3D PCAs

| ADE in HLT codes | PC1 (%) | PC2 (%) | PC3 (%) |
|---|---|---|---|
| Respiratory Failures (Excl Neonatal) | 8.02 | 2.36 | 0.00 |
| Respiratory Tract Disorders Nec | 7.81 | 1.53 | 0.01 |
| Bronchospasm and Obstruction | 7.80 | 0.92 | 0.94 |
| Parenchymal Lung Disorders Nec | 7.67 | 1.75 | 0.00 |
| Pulmonary Oedemas | 7.62 | 1.45 | 0.04 |
| Coughing and Associated Symptoms | 7.44 | 1.12 | 0.00 |
| Pneumothorax and Pleural Effusions Nec | 7.42 | 1.09 | 0.02 |
| Pulmonary Thrombotic and Embolic Conditions | 7.32 | 0.85 | 0.02 |
| Breathing Abnormalities | 7.15 | 0.62 | 0.01 |
| Lower Respiratory Tract Inflammatory and Immunologic Conditions | 6.89 | 0.32 | 0.03 |
| Lower Respiratory Tract Signs and Symptoms | 6.58 | 0.01 | 0.07 |
| Lower Respiratory Tract Infections Nec | 6.33 | 0.00 | 0.08 |
| Respiratory Signs and Symptoms Nec | 2.56 | 5.92 | 3.73 |
| Pleural Infections and Inflammations | 2.54 | 13.09 | 0.65 |
| Vascular Pulmonary Disorders Nec | 2.50 | 13.94 | 0.01 |
| Bronchial Conditions Nec | 2.45 | 10.83 | 0.64 |
| Respiratory Syncytial Viral Infections | 1.06 | 13.88 | 4.99 |
| Fungal Lower Respiratory Tract Infections | 0.41 | 10.39 | 63.19 |
| Pleural Conditions Nec | 0.36 | 14.84 | 0.87 |
| Lower Respiratory Tract Neoplasms | 0.04 | 2.44 | 11.18 |
| Infectious Disorders Carrier | 0.02 | 1.96 | 3.82 |
| Occupational Parenchymal Lung Disorders | 0.01 | 0.03 | 0.09 |
| Respiratory Tract Infections Nec | 0.00 | 0.01 | 9.33 |
| Pleural Neoplasms | 0.00 | 0.00 | 0.03 |
| Bacterial Lower Respiratory Tract Infections | 0.00 | 0.67 | 0.21 |

**PCA when excluding antihypertensive drugs:**

**(A)**

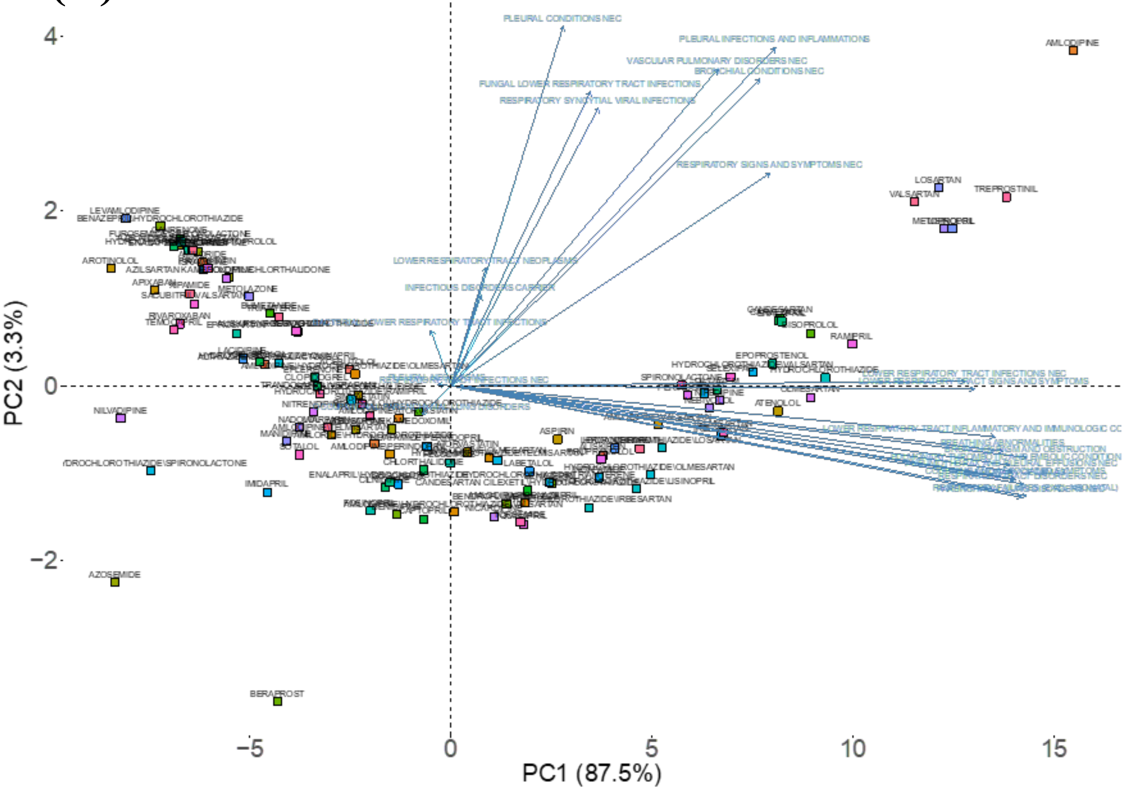


**(B)**

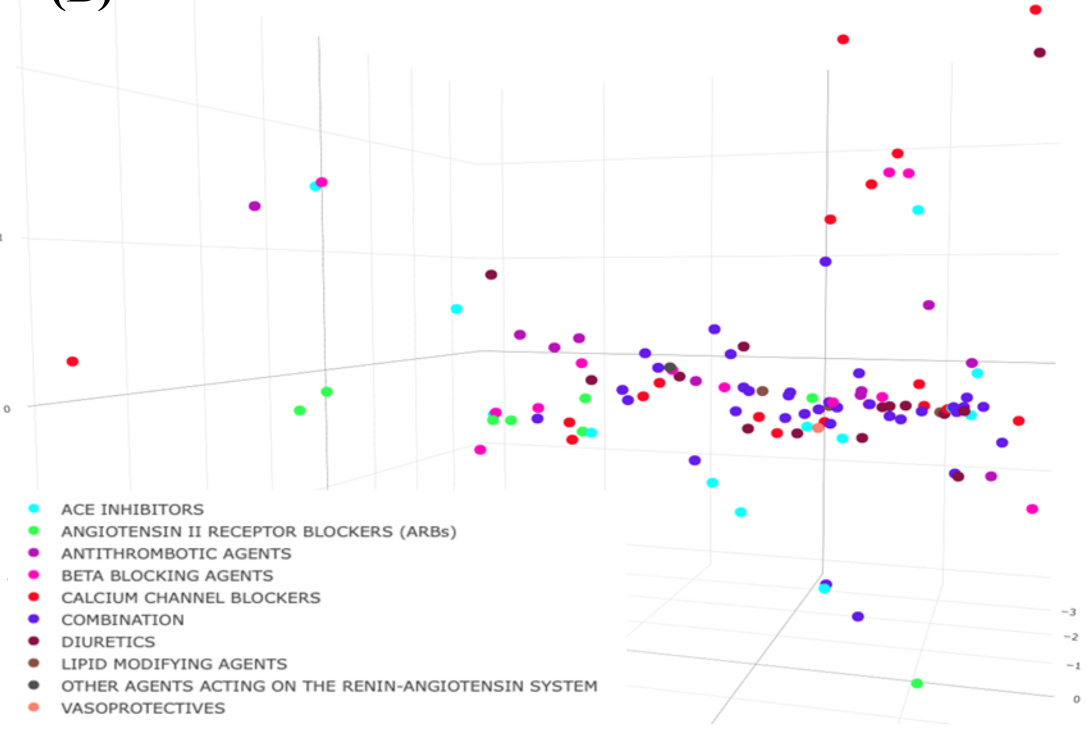

**Fig S1**. **Principal component analysis of expected count for all the 117 drugs** in 2D (A) and 3D (B) space using EBGM for ACEIs, ARBs, BBAs, CCBs, and TDs. In Panel B, individual drugs are (significantly) separated on the extreme edges are marked by (1) amlodipine, (2) quinapril, (3) trandolapril, (4) nilvadipine, (5) azosemide, (6) azelnidipine, and (7) Treprostinil

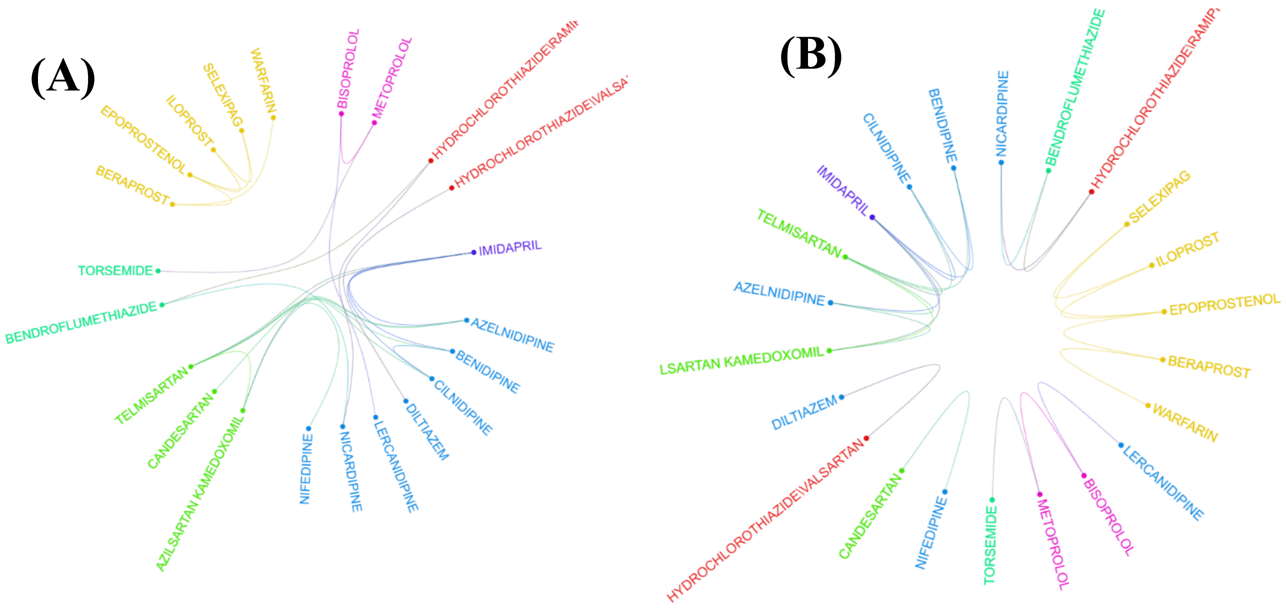


**Fig S2. Two layouts of Circos plot for 22 hypertensive drugs selected by GLASSO when excluding AHAs.** Circos plots of drugs were obtained based on the EBGM matrix after applying GLASSO. Edge bundling linkages for better visualization and drugs were selected by GLASSO with edge bundling. Grouped drugs in high-level terms were assigned the same color based on their classes (A). Applying RCM reordering and edge bundling for grouping drugs based on the ATC class and edge bundling (B).

**Table S2.** Frequency of pulmonary ADEs when RR larger than two or the 5th quantile of EBGM, EB05, large than two

| Drug | Pul. Count | Order by RR | Pul. Count | Order by EBGM |
|---|---|---|---|---|
| ILOPROST | 10 | 2 | 13 | 1 |
| EPOPROSTENOL | 9 | 4 | 11 | 2 |
| SELEXIPAG | 10 | 3 | 11 | 3 |
| BERAPROST | 12 | 1 | 7 | 5 |
| CANDESARTAN | 3 | 33 | 5 | 9 |
| NIFEDIPINE | 5 | 15 | 5 | 10 |
| WARFARIN | 5 | 17 | 5 | 12 |
| BISOPROLOL | N/A | N/A | 4 | 13 |
| DILTIAZEM | 1 | 73 | 3 | 16 |
| HYDROCHLOROTHIAZIDE\RAMIPRIL | 5 | 13 | 3 | 17 |
| IMIDAPRIL | 5 | 14 | 3 | 18 |
| LERCANIDIPINE | 1 | 83 | 3 | 20 |
| NICARDIPINE | 4 | 27 | 3 | 21 |
| TELMISARTAN | 4 | 28 | 3 | 23 |
| TORSEMIDE | 5 | 16 | 3 | 24 |
| AZELNIDIPINE | 4 | 22 | 2 | 26 |
| AZILSARTAN KAMEDOXOMIL | 3 | 31 | 2 | 27 |
| BENDROFLUMETHIAZIDE | 3 | 32 | 2 | 28 |
| BENIDIPINE | 5 | 9 | 2 | 29 |
| CILNIDIPINE | 5 | 11 | 2 | 30 |
| HYDROCHLOROTHIAZIDE\VALSARTAN | 1 | 80 | 2 | 33 |
| METOPROLOL | 1 | 86 | 2 | 34 |

**Table S3.** Comparative analysis of drug and associated pulmonary ADEs in different GLASSO Clusters

| Drug | Drug Class | ADEs for EB05> 1 (n) * | GL Cluster |
|---|---|---|---|
| ILOPROST | ATAs | 2,4-13,16-17 (13) | 1 |
| EPOPROSTENOL | ATAs | 1-2,4,7-13,16 (11) | 1 |
| SELEXIPAG | ATAs | 1-2,4-5,7-8,10-13,16 (11) | 1 |
| BERAPROST | ATAs | 1-2,4,10-13 (7) | 1 |
| WARFARIN | ATAs | 1,4,8,10,13 (5) | 1 |
| CANDESARTAN | ARBs | 1-3,6,15 (5) | 2 |
| NIFEDIPINE | CCBs | 1-3,9,15 (5) | 2 |
| BISOPROLOL | BBAs | 1-2,4,6 (4) | 3 |
| LERCANIDIPINE | CCBs | 1,4,6 (3) | 3 |
| TORSEMIDE | TDAs | 2-3,6 (3) | 3 |
| METOPROLOL | BBAs | 2,6 (2) | 3 |
| DILTIAZEM | CCBs | 2,5,9 (3) | 4 |
| HYDROCHLOROTHIAZIDE\VALSARTAN | COMBs | 5,9 (2) | 4 |
| IMIDAPRIL | ACEs | 1-3 (3) | 5 |
| TELMISARTAN | ARBs | 1-3 (3) | 5 |
| AZELNIDIPINE | CCBs | 1,3 (2) | 5 |
| AZILSARTAN KAMEDOXOMIL | ARBs | 1,3 (2) | 5 |
| BENIDIPINE | CCBs | 1-2 (2) | 5 |
| CILNIDIPINE | CCBs | 1-2 (2) | 5 |
| HYDROCHLOROTHIAZIDE\RAMIPRIL | COMBs | 2,7,14 (3) | 6 |
| NICARDIPINE | CCBs | 3,7,14 (3) | 6 |
| BENDROFLUMETHIAZIDE | TDAs | 3,14 (2) | 6 |

* Below we have ADEs found for each drug:
1. PARENCHYMAL LUNG DISORDERS NEC
2. PNEUMOTHORAX AND PLEURAL EFFUSIONS NEC
3. LOWER RESPIRATORY TRACT INFLAMMATORY AND IMMUNOLOGIC CONDITIONS
4. PULMONARY OEDEMAS
5. BRONCHOSPASM AND OBSTRUCTION
6. PULMONARY THROMBOTIC AND EMBOLIC CONDITIONS
7. RESPIRATORY TRACT DISORDERS NEC
8. COUGHING AND ASSOCIATED SYMPTOMS
9. LOWER RESPIRATORY TRACT INFECTIONS NEC
10. PARENCHYMAL LUNG DISORDERS NEC
11. PNEUMOTHORAX AND PLEURAL EFFUSIONS NEC
12. LOWER RESPIRATORY TRACT INFLAMMATORY AND IMMUNOLOGIC CONDITIONS
13. PULMONARY OEDEMAS
14. BRONCHOSPASM AND OBSTRUCTION
15. PULMONARY THROMBOTIC AND EMBOLIC CONDITIONS
16. RESPIRATORY TRACT DISORDERS NEC
17. COUGHING AND ASSOCIATED SYMPTOMS

**Arc Diagram**

Here we introduced a new data visualization method, called arc diagram, which is accomplished by representing complex patterns of repetition in the analysis of ADE-drug combination data. The arc diagram improves over previous methods because they efficiently scale for a drug-ADE combination that detects many instances and signals of the same repeated sub-signals. **Fig S3** and its zoom level inset maps describe design and implementation related to parallel arc diagrams of ADE-drug combination and show how they may be applied to visualize such diverse data and compiled code. This arc diagram consists of selected drugs from GLASSO and pulmonary-related ADEs as the vertices, and arches denote the associations of either drug-drug or drug-ADE. Here we have highlighted three inset maps as examples of these highly correlated and complex interactions. The association regarding drug-drug and drug-ADE selected by GLASSO method for $EB05 > 1$ was illustrated in an arc diagram (**Fig S3**). AHAs (macitentan, and bosentan) were found to have more pulmonary ADEs than other hypertensive drug classes. Two ATA drugs (epoprostenol and selexipag) showed more than 10 pulmonary ADEs. Given the clusters from GLASSO, all drugs except for group 1 had equal or less than five pulmonary ADEs. GLASSO group 1 consists of seven drugs from three ATC classes (from Table 3: AHAs, UAs, and ATAs). Combining RCM and arc diagram provides a more distinct way to demonstrate the association between drugs and their correlated pulmonary ADEs. The severity of pulmonary events associated with macitentan, bosentan, epoprostenol, selexipag, sildenafil, tadalafil, and beraprost was higher than the remaining drugs regarding six common ADEs in all of them. However, no more than two pulmonary issues were identical among drugs from the same GL Clusters 3, 4, 5, and 6, **Table 3**.

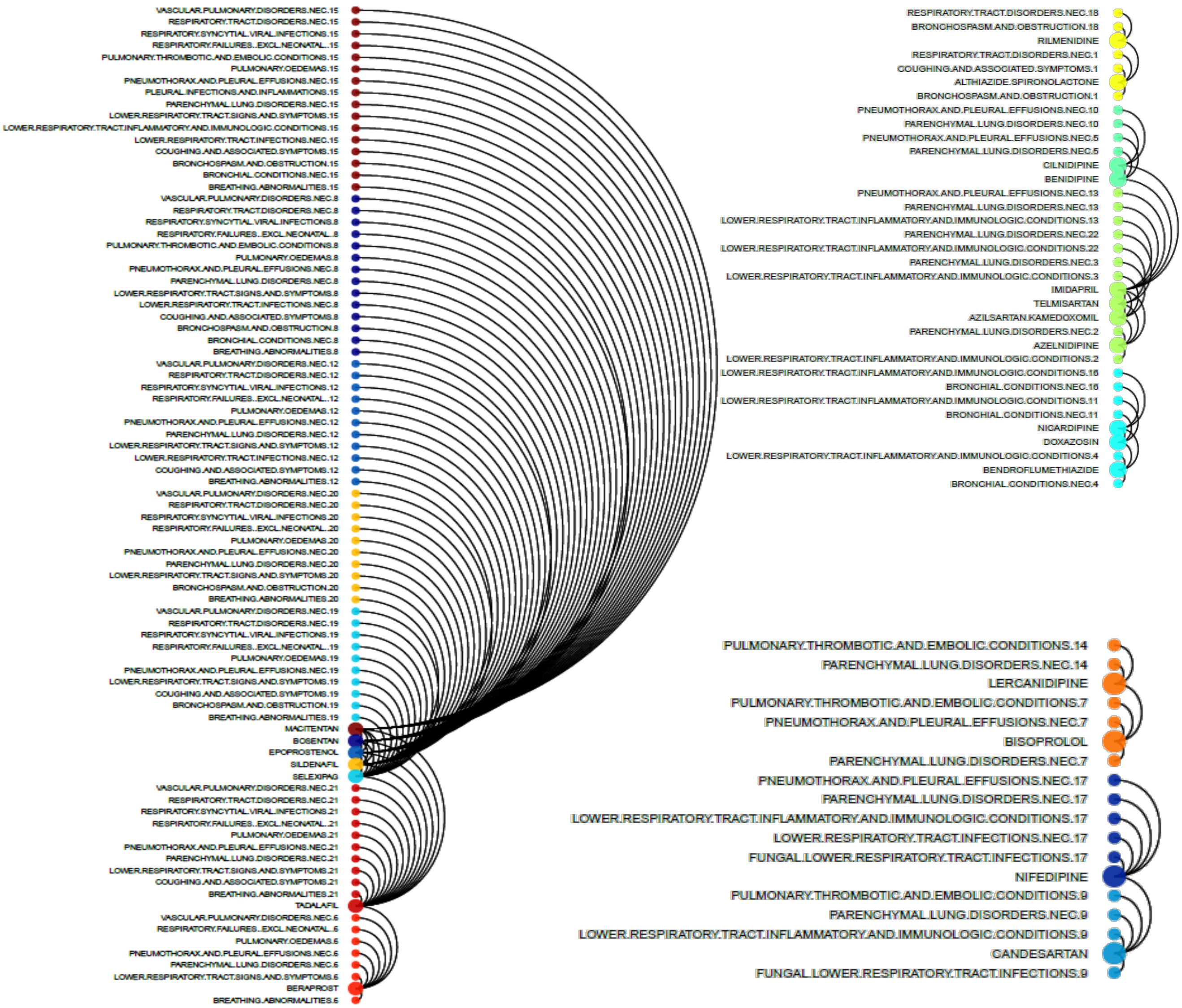


**Fig S3. Arc Diagram Visualization of 22 drugs selected from GLASSO and associated pulmonary ADE-drug combination.** Three inset maps as examples of highly correlated and complex interactions are shown. Large-filled circles show drugs and small circles are used for ADEs.

**Table S4.** Friedman test for drugs in ATC class and GLASSO class

| ATC Class | p-value (44 drugs) | p-value (22 drugs) |
|---|---|---|
| ACEIs | 0.229 | - |
| ARBs | **0.0056** | **0.0129** |
| ATAs | **< 0.001** | **0.007** |
| BBAs | **0.007** | 1 |
| CCBs | **0.00477** | 0.095 |
| COMBs | 0.369 | 0.819 |
| TDAs | 0.0552 | 0.108 |

| GL Cluster | p-value for 22 drugs |
|---|---|
| 1 | **0.00736** (0.196, when excluding Warfarin) |
| 2 | 0.127 |
| 3 | 0.647 |
| 4 | 0.0736 |
| 5 | 0.0675 |
| 6 | 0.0840 |

**Table S5-A.** Multiple comparisons of different ATC classes together with the adjusted p-value using the rigorous paired Wilcoxon signed-rank test with Bonferroni correction.

| ATC Group | ATC Group | p-value | Adj. p-value | ATC Group | ATC Group | p-value | Adj. p-value |
|---|---|---|---|---|---|---|---|
| ACEIs | ARBs | 0.33 | 1 | **ATAs** | BBAs | **0.009** | 0.341 |
| ACEIs | ATAs | **0.021** | 0.763 | **ATAs** | CCBs | **0.006** | 0.234 |
| ACEIs | ATAs | **0.002** | 0.066 | **ATAs** | COMBs | **0.002** | 0.084 |
| ACEIs | BBAs | 0.267 | 1 | **ATAs** | TDAs | **0.015** | 0.54 |
| ACEIs | CCBs | 0.475 | 1 | **ATAs** | UAs | **0.006** | 0.217 |
| ACEIs | COMBs | 0.977 | 1 | **BBAs** | CCBs | 0.727 | 1 |
| ACEIs | TDAs | 1 | 1 | **BBAs** | COMBs | 0.433 | 1 |
| ACEIs | UAs | **0.024** | 0.86 | **BBAs** | TDAs | 0.401 | 1 |
| ARBs | AHAs | **0.043** | 1 | **BBAs** | UAs | **0.042** | 1 |
| ARBs | AHAs | **0.013** | 0.475 | **CCBs** | COMBs | 0.436 | 1 |
| ARBs | BBAs | 0.603 | 1 | **CCBs** | TDAs | 0.626 | 1 |
| ARBs | CCBs | 0.867 | 1 | **CCBs** | UAs | **0.027** | 0.986 |
| ARBs | COMBs | 0.495 | 1 | **COMBs** | TDAs | 0.698 | 1 |
| ARBs | TDAs | 0.455 | 1 | **COMBs** | UAs | **0.022** | 0.785 |
| ARBs | UAs | 0.066 | 1 | **TDAs** | UAs | 0.065 | 1 |
| AHAs | ATAs | **0.015** | 0.536 | | | | |
| AHAs | BBAs | **0.034** | 1 | | | | |
| AHAs | CCBs | **0.016** | 0.562 | | | | |
| AHAs | COMBs | **0.009** | 0.338 | | | | |
| AHAs | TDAs | **0.03** | 1 | | | | |
| AHAs | UAs | 0.399 | 1 | | | | |

**Table S5-B.** Multiple comparisons of different ATC classes

| ATC Group | ATC Group | p-value | Adj. p |
|---|---|---|---|
| ACEIs | ARBs | 0.506 | 1 |
| ACEIs | ATAs | **0.005** | 0.102 |

| | | | |
|---|---|---|---|
| ACEIs | BBAs | 0.224 | 1 |
| ACEIs | CCBs | 0.354 | 1 |
| ACEIs | COMBs | 0.421 | 1 |
| ACEIs | TDAs | 0.344 | 1 |
| ARBs | ATAs | **0.017** | 0.357 |
| ARBs | BBAs | 0.494 | 1 |
| ARBs | CCBs | 0.449 | 1 |
| ARBs | COMBs | 0.745 | 1 |
| ARBs | TDAs | 0.399 | 1 |
| ATAs | BBAs | **0.011** | 0.237 |
| ATAs | CCBs | **0.014** | 0.284 |
| ATAs | COMBs | **0.011** | 0.235 |
| ATAs | TDAs | **0.016** | 0.342 |
| BBAs | CCBs | 0.966 | 1 |
| BBAs | COMBs | 0.495 | 1 |
| BBAs | TDAs | 0.702 | 1 |
| CCBs | COMBs | 0.704 | 1 |
| CCBs | TDAs | 0.637 | 1 |
| COMBs | TDAs | 0.723 | 1 |

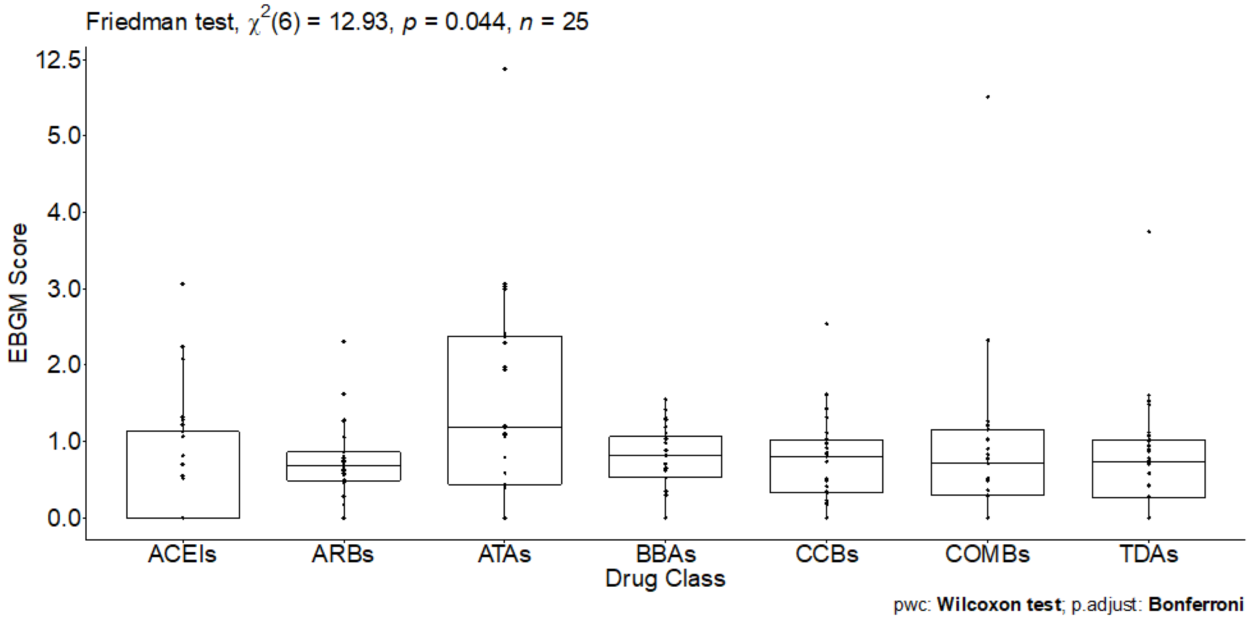


**Fig S4**. **Pairwise Wilcoxon signed-rank test between different ATC classes.** No pairwise significant comparison was found similar to **Table S5-B**. But the group comparison was very significant, *p-value=0.044*.

**Table S6-A.** Multiple comparisons of drugs from GL Clusters and multiple comparisons of drugs from GL clusters excluding Tadalafil.

| GL Cluster | GL Cluster | Including Tadalafil | | Excluding Tadalafil | |
|---|---|---|---|---|---|
| | | p-value | Adj. p-value | p-value | Adj. p-value |
| 1 | 2 | **0.038** | 0.566 | **0.027** | 0.41 |
| 1 | 3 | **0.000579** | **0.009**** | **0.000355** | **0.005**** |
| 1 | 4 | **0.002** | **0.034*** | **0.000928** | **0.014*** |
| 1 | 5 | **0.004** | 0.056 | **0.002** | **0.024*** |
| 1 | 6 | **0.007** | 0.106 | **0.004** | 0.064 |
| 2 | 3 | 0.052 | 0.774 | 0.052 | 0.774 |
| 2 | 4 | 0.135 | 1 | 0.135 | 1 |
| 2 | 5 | **0.037** | 0.558 | **0.037** | 0.558 |
| 2 | 6 | 0.056 | 0.839 | 0.056 | 0.839 |
| 3 | 4 | 0.142 | 1 | 0.142 | 1 |
| 3 | 5 | 0.424 | 1 | 0.424 | 1 |
| 3 | 6 | 0.191 | 1 | 0.191 | 1 |
| 4 | 5 | 0.354 | 1 | 0.354 | 1 |
| 4 | 6 | 0.972 | 1 | 0.972 | 1 |
| 5 | 6 | 0.475 | 1 | 0.475 | 1 |

**Table S6-B.** Multiple comparisons of drugs from GLASSO clusters and multiple comparisons of drugs from GLASSO clusters excluding Warfarin.

| GL Cluster | GL Cluster | Including Warfarin | | Excluding Warfarin | |
|---|---|---|---|---|---|
| | | p-value | Adj. p-value | p-value | Adj. p-value |
| 1 | 2 | **0.027** | 0.41 | **0.012** | 0.186 |
| 1 | 3 | **0.008** | 0.116 | **0.004** | 0.058 |
| 1 | 4 | **0.027** | 0.41 | **0.012** | 0.186 |
| 1 | 5 | **0.006** | 0.095 | **0.003** | **0.047*** |
| 1 | 6 | **0.005** | 0.079 | **0.003** | **0.042*** |
| 2 | 3 | 0.205 | 1 | 0.205 | 1 |
| 2 | 4 | 0.559 | 1 | 0.559 | 1 |
| 2 | 5 | **0.05** | 0.747 | **0.05** | 0.747 |
| 2 | 6 | 0.154 | 1 | 0.154 | 1 |
| 3 | 4 | 0.884 | 1 | 0.884 | 1 |
| 3 | 5 | 0.204 | 1 | 0.204 | 1 |
| 3 | 6 | 0.639 | 1 | 0.639 | 1 |
| 4 | 5 | 0.176 | 1 | 0.176 | 1 |
| 4 | 6 | 0.537 | 1 | 0.537 | 1 |
| 5 | 6 | 0.516 | 1 | 0.516 | 1 |

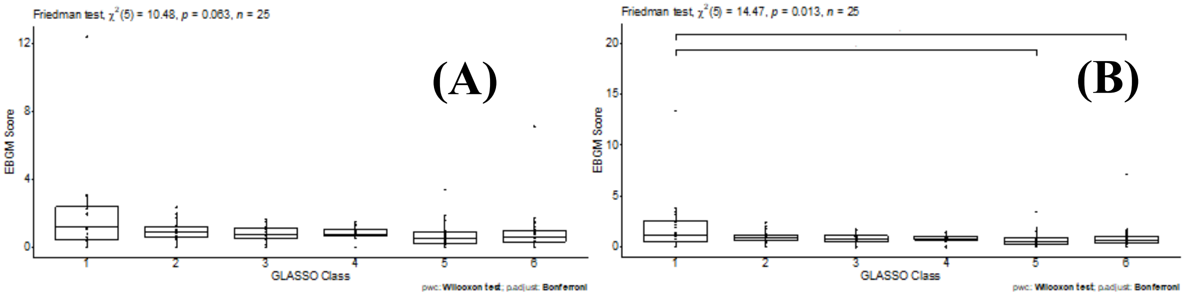


**Fig S5. Pairwise Wilcoxon signed-rank test for different classes** defined by GLASSO (A) and the same test for different classes defined by GLASSO excluding Warfarin (B) similar to **Table S6-B**.

**Table S7.** Dose distribution related to tadalafil and sildenafil and ADE.

| Tadalafil Dose | Frequency | Sildenafil Dose | Frequency |
|---|---|---|---|
| 40 | 5270 | 100 | 5997 |
| 20 | 5024 | 60 | 5775 |
| 5 | 3686 | 50 | 2913 |
| 1 | 1784 | 20 | 1647 |
| 10 | 1242 | 25 | 927 |
| 2.5 | 455 | 120 | 845 |
| 2 | 220 | 40 | 606 |
| 80 | 74 | 1 | 563 |
| 60 | 63 | 240 | 335 |
| 30 | 58 | 75 | 322 |
| 0.5 | 53 | 150 | 300 |
| 15 | 48 | 200 | 297 |
| 100 | 26 | 180 | 273 |
| 25 | 20 | 3 | 248 |
| 4 | 17 | 30 | 230 |
| 120 | 17 | 80 | 178 |
| 3 | 16 | 10 | 159 |
| 50 | 15 | 2 | 152 |
| 7.5 | 13 | 0.5 | 149 |
| 8 | 8 | 300 | 142 |
| 400 | 6 | 6 | 88 |
| 200 | 6 | 5 | 65 |
| 1.25 | 6 | 4 | 57 |
| 250 | 5 | 90 | 42 |
| 22 | 5 | 9 | 42 |
| 7 | 4 | 15 | 40 |
| 6 | 4 | 400 | 31 |
| 160 | 4 | 225 | 31 |
